## Supplemental material for "The epidemiology of Mayaro virus in the Americas: A systematic review and key parameter estimates for outbreak modelling"

**Supporting S1 Text**

Edgar-Yaset Caicedo^1^, Kelly Charniga^2^, Amanecer Rueda^1^, Ilaria Dorigatti^2^, Yardany Mendez^1^, Arran Hamlet^2^, Jean-Paul Carrera^3,4^, Zulma M. Cucunubá^2*^

^1^Universidad Pedagógica y Tecnológica de Colombia, Tunja, Colombia

^2^MRC Centre for Global Infectious Disease Analysis (MRC-GIDA), Imperial College London, London, UK

^3^Department of Zoology, University of Oxford, Oxford, UK

^4^Department of Research in Virology and Biotechnology, Gorgas Memorial Institute of Health Studies, Panama City, Panamá

+ These authors contributed equality to this work

**Supplementary methods for systematic review**

Records identified by searching databases (n = 1222)

Records after duplicates removed

(n = 713)

Records screened

(n = 713)

Unrelated records excluded

(n = 573)

Full-text articles assessed for eligibility

(n = 140)

Full text excluded because inclusion criteria not met (n = 50)

Studies included in analysis

(n = 76)

Identification

Screening

Eligibility

Included

**S1 Fig. Flowchart showing the selection of studies.**

**S1 Table. Boolean algorithms for literature search.**

| Database | Algorithm* | Number of titles | Dates |
| --- | --- | --- | --- |
| Web of knowledge | ((mayaro virus OR MAYV OR uruma)) | 168 | 11 Jan 2019 |
| PubMed | ((mayaro[All Fields] AND ("viruses"[MeSH Terms] OR "viruses"[All Fields] OR "virus"[All Fields])) OR uruma[All Fields]) OR MAYV[All Fields] | 434 | 11 Jan 2019 |
| LILACS | (tw:(mayaro virus OR fiebre de mayaro OR MAYV OR uruma)) | 248 | 11 Jan 2019 |
| EMBASE | mayaro virus.af  MAYV.af  uruma.ab  #1 OR #2 OR #3 | 213 | 15 Jan 2019 |
| Google  Scholar | (Mayaro Virus OR MAYV) | 159 | 11 Jan 2019 |
| Total |  | 1222 |  |

*No restrictions were used on any of the search terms.

**S2 Table. Data classification of MAYV studies in humans.**

| Study type | Definition |
| --- | --- |
| Case reports | A detailed report describing one or more confirmed cases.  Tests: polymerase chain reaction (PCR), IgM, or isolates or a combination of these with symptom surveillance, or seroconversion by IgG (with evidence of previous IgG negative), or an increased titre > 4 times the previous sample. |
| Outbreaks | An outbreak of confirmed and suspected cases is reported in detail (location, period of time, etc).  Tests: PCR, IgM, or isolates or a combination of these with symptom surveillance or seroconversion by IgG (with evidence of previous IgG negative) or an increased titre > 4 times to previous sample, with or without reported symptoms. |
| Hospital-based surveillance | A study that is performed in a health facility with symptomatic patients.  Tests: PCR, IgM (HI, ELISA), or isolates or a combination of these with symptom surveillance or seroconversion by IgG (HI, ELISA, complement fixation), (with evidence of previous IgG negative) or an increased titre of > 4 times the previous sample. IgG by ELISA or HI only if the samples tested negative against all other alphaviruses’ antigens. The health facilities were classified according to their location into urban, rural, and unknown location. |
| Cross-sectional seroprevalence | A study conducted in a community of asymptomatic people that is representative of the general population of that community.  Tests: antibodies against MAYV using one or more of the following techniques: platelet reaction neutralisation test (PRNT), neutralisation test (NT), ELISA or hemagglutination inhibition (HI). |
| Others | Not classified in other categories but strongly indicates MAYV presence. |

**Supplementary methods for generation time and time-varying reproduction number**

**Natural history parameter estimates**

**Viral load data.** We found one paper that reported the absolute quantification of Mayaro RNA viral copies in plasma following the onset of symptoms [1]. This paper reported the duration of viremia in 21 patients from whom Mayaro virus (MAYV) was isolated. Viremia data was reported on the log_10_ scale and presented as mean values and ranges (S2 Fig).

Mayaro viral loads in humans are comparable to those of Zika [2].

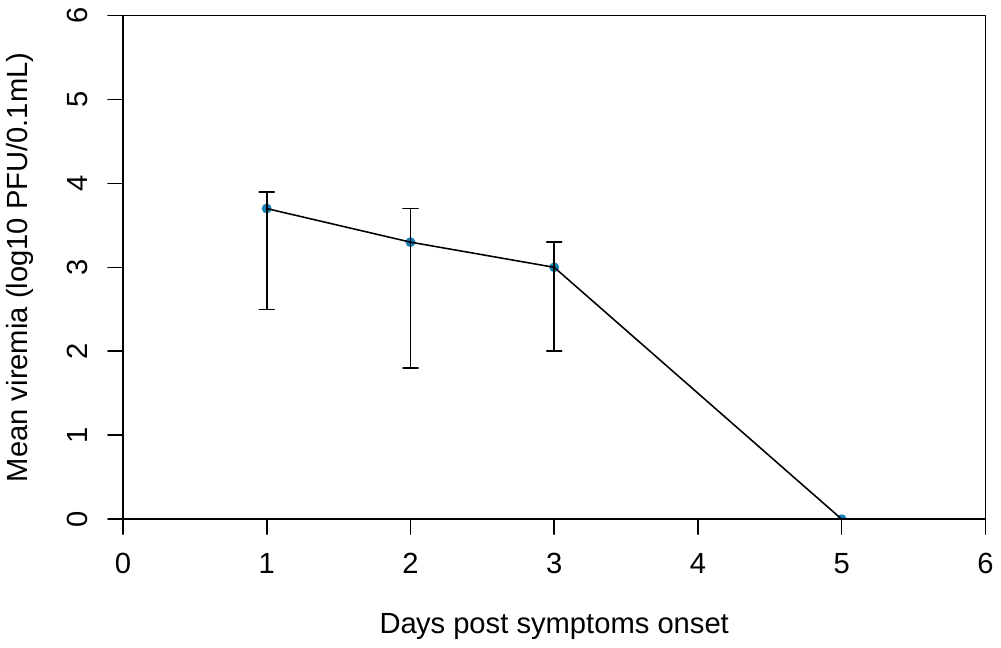

**S2 Fig. Viral load detected in plasma in Mayaro infected cases per day post symptoms onset.** Mean and range for 21 patients are shown. Adapted from [1].

**Human to mosquito generation time.** The human-to-mosquito generation time is the time between human infection and a mosquito taking an infectious blood meal. It is composed of the intrinsic incubation period (the time between human infection and symptoms onset) and the time from symptoms onset to viral clearance.

**Intrinsic incubation period.** We found 10 peer-reviewed articles that reported information on time of exposure to MAYV and time of symptoms onset in humans. From these, we extracted data on 15 cases of MAYV. All were infected while traveling to endemic areas. Most cases were infected after 2004, no cases were reported in children, and of the 11 studies that reported the sex of patients, six were males and five were females.

We excluded articles if they did not report quantitative information on time of exposure and symptom onset. We used the exact timing of exposure whenever possible. When this information was not reported, we used the information provided to bound the time of exposure. Following, we bounded the time of MAYV infection by the earliest and latest potential times of exposure [3].

A doubly interval censored dataset was constructed for the incubation period and the distribution was fitted using the methods described in [3, 4]. Following Lessler et al. [3], we assumed that the incubation period of MAYV followed a log-normal distribution and used the Metropolis-Hastings Markov chain Monte Carlo (MCMC) algorithm for calibration.

We estimated a mean incubation period μ_IP_ of 3.0 (95% CrI: 2.2-3.8) days and a standard deviation σ_IP_ of 1.2 (95% CrI: 1.0-1.7) days.

**Time to viral clearance.** Following [2], we assumed the time to viral clearance is Gamma distributed with shape parameter $\alpha_{C}$ and scale parameter $\beta_{C}$. We estimated $\alpha_{C}$ and $\beta_{C}$ using the Metropolis-Hastings MCMC algorithm. In Table 3 of the main text we report the mean ($\mu_{C}=\alpha_{C}\beta_{C}$) and standard deviation ($\sigma_{C}=\alpha_{C}\beta_{C}^{2}$) of the time to viral clearance.

We also assumed that infectiousness in Mayaro infection is similar to Zika infection, beginning 1.5 days before symptom onset and ending 1.5-2 days before viral levels can no longer be detected. To obtain the human generation time from the intrinsic incubation period and time to viral clearance, we linearly scaled the time dependence of the distribution by a factor $s=\left( \mu_{IP}-1.5 \right)/\mu_{IP}$. This results in a human generation time with mean $\mu_{h}=s\left( \mu_{IP}+\mu_{C} \right)$ and standard deviation $\sigma_{h}=s\sqrt{\sigma_{IP}^{2}+\sigma_{C}^{2}}$.

**Extrinsic incubation period.** We found five peer-reviewed articles that reported the susceptibility of mosquitoes to MAYV [5-9]. Only three articles [6, 8, 9] reported enough information on the number of mosquitoes tested at each day post-infection, which are necessary to estimate the extrinsic incubation period. Across these studies, seven different species of mosquitoes and three different strains of MAYV were used. Due to limited data, we combined information across all species of mosquitoes and viral strains. We dropped *Cx. quinquefasciatus* from the pooled analysis because it had null transmission rates. We also dropped observations on day 14 from [6] because the observed frequency did not match with the pattern observed in the rest of the data. We defined the extrinsic incubation period as the time between infection and MAYV reaching the salivary glands.

As in [2], we assumed that the extrinsic incubation period is Gamma distributed with shape parameter k_EIP_ and scale parameter θ_EIP_. Using a Binomial likelihood function to estimate the probability that a mosquito is infectious by day *t*, we obtain mean posterior estimates of k_EIP_ = 4.5 (95% CrI: 2.4-7.3) and θ_EIP_ = 2.3 (95% CrI: 1.2-4.3). This results in a mean EIP of 9.4 (95% CrI: 8.4-10.7) days with a standard deviation of 4.6 (95% CrI: 3.3-6.7) days (main text Table 3). S3 Fig shows the fitted probability density function, cumulative distribution function, and observed data.

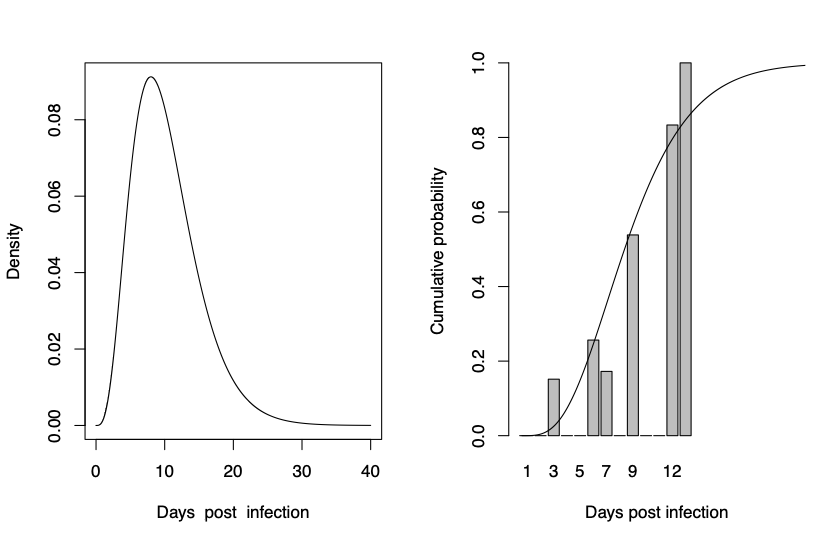

**S3 Fig.** **Maximum likelihood EIP probability density function (left) and cumulative distribution function (right).** The aggregated proportion of mosquitoes that tested positive at the relative days post-infection are shown as bars.

**Mosquito-to-human generation time.** We use the same methods as [2] to estimate the mosquito-to-human generation time (the time between a mosquito being infected and it infecting a human) from the estimated EIP and the mosquito daily mortality rate. As in [2], we assume that the mosquito mortality rate $ɛ$ is Gamma distributed with a mean of 0.2/day and a standard deviation of 0.05/day.

We define the probability density function of the mosquito-to-human generation time as

$$h\left( t \right)= \frac{f^{'}\left( t \right)e^{-ɛt}}{\int_{0}^{\infty} f^{'}\left( t^{'} \right)e^{-ɛt^{'}}dt'}$$

where $f'\left( t \right)$represents the density function of the EIP (the probability density of a mosquito being infectious at time $t$). We estimate the mean 𝜇_m_ and standard deviation σ_m_ of the mosquito-to-human generation time numerically, i.e. sampling the shape parameter k_EIP_ and scale parameter θ_EIP_ from their posterior distributions and $ɛ$ from the Gamma distribution with mean of 0.2/day and standard deviation of 0.05/day.

We estimated a mean mosquito-to-human generation time of 11.9 (95% CrI: 8.6 - 16.3) days and a standard deviation of 6.2 (95% CrI: 4.2 - 9.5) days (main text Table 3).

**Generation time of MAYV.** Combining the estimates of the human-to-mosquito generation time with those of the mosquito-to-human generation time, we estimate that the distribution of the generation time of MAYV (i.e. the time between infection of a human case and infection of the secondary human cases that case causes) has a mean of 15.2 (95% CrI: 11.7 - 19.8) days and a standard deviation of 6.2 (95% CrI: 4.2 - 9.5) days (main text Table 3).

**Estimates of the reproduction number, R**

We estimated the instantaneous reproduction number *R* for the 1954-1955 MAYV outbreak in Santa Cruz, Bolivia using the weekly case incidence and the generation time distribution estimated in the previous section.

The instantaneous reproduction number *R* was calculated over 4-week sliding windows using the EpiEstim package in R software [10]. The instantaneous reproduction number *R* of each time window is calculated as the median of the weekly instantaneous reproduction number weighted by the weekly incidence. *R* values were plotted in the middle of the 4-week time window used to calculate each estimate.

The analysis was performed in R (version 3.5.3). The *‘uncertain_si’* option was selected in the estimate_R() function of EpiEstim, using the mean and standard deviation of the generation time from Table 3 in the main text and a prior distribution for R with a mean and standard deviation of 5. All values can be found in S3 Table. The uncertain_si method takes into account uncertainty on the serial interval distribution as described in Cori et al. [11]. Briefly, the mean μ and standard deviation σ of the serial interval are permitted to vary according to truncated normal distributions. The values in S3 Table were used to sample n1 pairs of mean and standard deviations from their respective truncated normal distributions. For each pair, a sample of size n2 was drawn from the posterior distribution of the reproduction number over each time window. n2 was drawn conditionally on the serial interval distribution obtained. A sample of size n1xn2 of the joint posterior distribution of the reproductive number over each time window was generated after pooling.

**S3 Table. Values used in estimate_R() function in EpiEstim package.**

| Parameter | Days* |
| --- | --- |
| mean_si | 15.2 |
| std_mean_si | 2.1 |
| min_mean_si | 7.0 |
| max_mean_si | 29.5 |
| std_si | 6.3 |
| std_std_si | 1.3 |
| min_std_si | 3.0 |
| max_std_si | 17.0 |

*All values were divided by 7 for weekly data.

**Supplementary results for systematic review**

**S4 Table. Characteristics of MAYV case reports.**

| Ref | Year of detection | Country | State | Town/City | Diagnostic method* | Diagnostic certainty | Cases | Zone | Origin |
| --- | --- | --- | --- | --- | --- | --- | --- | --- | --- |
| [12] | 2012 | Bolivia | Beni | Rurrenabaque | IgM/IgG NT and IA | confirm | 1 | rural | foreign |
| [13] | 2000 | Brazil | Mato Grosso | Camapua | Culture, RT-PCR | confirmed | 3 | rural | foreign |
| [14] | 2004 | Brazil | Acre | Acrelandia | RT-PCR | confirmed | 1 | rural | native |
| [15] | 2009 | Brazil | Amazonas | Barcelos | IgM ELISA | potential | 1 | rural | foreign |
| [16] | 2013 | Brazil | Para |  | IgM | potential | 2 | rural | native |
| [17] | 2015 | Brazil | Sao Paulo | Sao Jose do Rio Preto | RT-PCR | confirmed | 1 | rural | foreign |
| [18] | 2015 | Brazil | Para | Portal | RT-PCR | confirmed | 1 | urban | native |
| [19] | 1996 | French Guiana |  |  | Culture, RT-PCR | confirmed | 1 | rural | native |
| [20] | 1998 | French Guiana | Cayenne | Cayenne | IgM ELISA | potential | 1 | urban | native |
| [21] | 2012 | French Guiana | Cayenne | Kourou | IgM ELISA | potential | 3 | mixed | native |
| [22] | 2013 | French Guiana |  |  | RT-PCR | potential | 1 | rural | foreign |
| [23] | 2015 | French Guiana | Cayenne | Roura | RT-PCR | confirmed | 1 | rural | foreign |
| [24] | 2014 | Haiti | Oest | Port-au-Prince | RT-PCR | confirmed | 1 | urban | native |
| [25] | 2001 | Mexico | Tamaulipas | Tampico | IgM ELISA | potential | 1 | urban | native |
| [25] | 2001 | Mexico | Veracruz | Coatzacoalco | IgM ELISA | potential | 1 | urban | native |
| [26] | 1995 | Peru | Loreto | Iquitos | Culture, IgM ELISA, PCR | confirmed | 2 | urban | foreign |
| [26] | 1995 | Peru | Loreto | Iquitos | Culture, IgM ELISA, PCR | confirmed | 13 | mixed | native |
| [26] | 1995 | Peru | Loreto | Yurimaguas | Culture, IgM ELISA, PCR | confirmed | 1 | mixed | native |
| [26] | 1995 | Peru | San Martin | Tocache | Culture, IgM ELISA, PCR | confirmed | 1 | mixed | native |
| [26] | 1995 | Peru | Ucayali | Pucallpa | Culture, IgM ELISA, PCR | confirmed | 6 | mixed | native |
| [26] | 1995 | Peru | Huanuco | Huanuco | Culture, IgM ELISA, PCR | confirmed | 3 | mixed | native |
| [26] | 1995 | Peru | Cusco | Quillabamba | Culture, IgM ELISA, PCR | confirmed | 1 | mixed | native |
| [26] | 1995 | Peru | Tumbes | Tumbes | Culture, IgM ELISA, PCR | confirmed | 2 | mixed | native |
| [27] | 2011 | Peru | San Martin | Tarapoto | IgM/IgG NT and IA | confirmed | 1 | rural | foreign |
| [28] | 2008 | Surinam |  |  | IgM ELISA | potential | 2 | rural | foreign |
| [29] | 1954 | Trinidad and Tobago | Mayaro | Cats Hill | Culture | confirmed | 5 | rural | native |
| [30] | 2000 | Venezuela | Miranda | Padron Agriculture | IgM ELISA | potential | 4 | rural | native |

NT: neutralization test, IA: immunofluorescence assay.

**S5 Table. Characteristics of Mayaro fever cases included in the intrinsic incubation period analysis (N = 15).**

| Ref | Age | Sex | Place of origin | Probable location infected | Year exposed | Exposure window (days) | Days to symptom onset (min-max) |
| --- | --- | --- | --- | --- | --- | --- | --- |
| [23] | 30 | Male | France | French Guiana | - | 6 | 1-8 |
| [22] | 44 | Female | Germany | French Guiana | 2013 | 18 | 1-20 |
| [16] | 52 | Female | Netherlands | Brazil | 2013 | 26 | 0-26 |
| [12] | 20 | Female | Germany | Bolivia | 2012 | 10 | 1-12 |
| [27] | 27 | Male | Switzerland | Peru | 2011 | 14 | 0-14 |
| [15] | Late 20s | Male | France | Brazil | 2010 | 14 | 0-14 |
| [13] | 36  59  74 | Male  Male  Male | SP, Brazil  SP, Brazil  SP, Brazil | MS, Brazil  MS, Brazil  MS, Brazil | 2000  2000  2000 | 8  9  8 | 0-8  0-10  0-8 |
| [30] | 26-58  26-58  26-58 | -  -  - | -  -  - | Venezuela  Venezuela  Venezuela | 2000  2000  2000 | 1  1  1 | 2-4  2-4  2-4 |
| [26] | 48  29 | Female  Female | USA  USA | Peru  Peru | 1996  1997 | 14  75 | 0-15  0-75 |
| [1] | - | - | Brazil | PA, Brazil | 1978 | 7 | 0-7 |

**S6 Table. Characteristics of hospital-based surveillance studies included in the analysis.**

| Ref | Study years | Country | State | Town/city | Diagnostic method* | Other arboviruses studied | Total | No. positive | % positive | Zone | Origin |
| --- | --- | --- | --- | --- | --- | --- | --- | --- | --- | --- | --- |
| [31] | 2000-2007 | Bolivia | Santa Cruz | Santa Cruz | Culture, RT PCR, IgM ELISA | VEEV, MURV, CARV, EEV, YFV, OROV, GUAV, DENV | 1280 | 10 | 0.8% | unknown | native |
| [31] | 2004-2006 | Bolivia | Beni | Magdalena | Culture, RT PCR, IgM ELISA | VEEV, MURV, CARV, EEV, YFV, OROV, GUAV, DENV | 173 | 1 | 0.6% | unknown | native |
| [31] | 2004-2007 | Bolivia | Santa Cruz | Concepcion | Culture, RT PCR, IgM ELISA | VEEV, MURV, CARV, EEV, YFV, OROV, GUAV, DENV | 380 | 3 | 0.8% | unknown | native |
| [31] | 2005-2007 | Bolivia | Cochabamba | Cochabamba | Culture, RT PCR, IgM ELISA | VEEV, MURV, CARV, EEV, YFV, OROV, GUAV, DENV | 256 | 10 | 3.9% | unknown | native |
| [32] | 1998-1999 | Brazil | Amazonas | Manaus | IgM ELISA | DENV, OROV | 8577 | 8 | 0.1% | urban | native |
| [33] | 2007-2008 | Brazil | Amazonas | Manaus | Culture, RT PCR, IgM ELISA | no | 631 | 33 | 5.2% | urban | native |
| [34] | 2009 | Brazil | Para | Novo Progresso | IgM ELISA and HI | DENV, YFV, OROV | 744 | 28 | 3.8% | unknown | native |
| [34] | 2009 | Brazil | Para | Trairão | IgM ELISA and HI | DENV, YFV, OROV | 654 | 49 | 7.5% | unknown | native |
| [35] | 2011-2013 | Brazil | Goias | Goiania | IgM EIA-ICC ELISA | DENV, OROV | 647 | 6 | 0.9% | unknown | native |
| [36] | 2011-2012 | Brazil | Mato Grosso | Sinop | RT-PCR | UNAV, SFV, Getah virus, Ross River virus | 200 | 6 | 3.0% | urban | native |
| [37] | 2011-2012 | Brazil | Mato Grosso |  | RT-PCR | AURV, WEEV, EEEV, DENV | 604 | 15 | 2.5% | unknown | native |
| [38] | 2014-2015 | Brazil | Goias |  | IgM MAC-ELISA and HI | DENV, CHIKV | 75 | 15 | 20.0% | unknown | foreign |
| [39] | 2015-2016 | Brazil | Mato Grosso |  | RT-PCR | DENV, YF, SLEV, ILHV, ROCV, WNV, EEEV, WEEV, VEEV, CHIKV | 453 | 34 | 7.5% | unknown | native |
| [40] | 2016-2017 | Brazil | Piaui | Parnaiba | RT-PCR | DENV, CHIKV | 578 | 1 | 0.2% | urban | native |
| [31] | 2003-2007 | Ecuador | Guayas | Guayaquil | Culture, RT PCR, IgM ELISA | VEEV, MURV, CARV, EEV, YFV, OROV, GUAV, DENV | 350 | 1 | 0.3% | unknown | native |
| [41] | 2003-2016 | French Guiana | Cayenne |  | RT-PCR or IgM ELISA | no | 412 | 9 | 2.2% | urban | native |
| [31] | 2000-2007 | Peru | Cusco | Cusco | Culture, RT PCR, IgM ELISA | VEEV, MURV, CARV, EEV, YFV, OROV, GUAV, DENV | 826 | 4 | 0.5% | unknown | native |
| [42] | 2000-2001 | Peru | Morropon | Salitral | IgM-ELISA | DENV, YFV, OROV, VEEV | 65 | 1 | 1.5% | unknown | native |
| [31] | 2004-2007 | Peru | Loreto | Yurimaguas | Culture, RT PCR, IgM ELISA | VEEV, MURV, CARV, EEV, YFV, OROV, GUAV, DENV | 1452 | 11 | 0.8% | unknown | native |
| [31] | 2004-2007 | Peru | Loreto | Iquitos | Culture, RT PCR, IgM ELISA | VEEV, MURV, CARV, EEV, YFV, OROV, GUAV, DENV | 10739 | 48 | 0.5% | unknown | native |
| [31] | 2004-2007 | Peru | Madre de Dios | Puerto Maldonado | Culture, RT PCR, IgM ELISA | VEEV, MURV, CARV, EEV, YFV, OROV, GUAV, DENV | 1215 | 10 | 0.8% | unknown | native |
| [43] | 2010-2013 | Peru | Loreto | Iquitos | Culture, RT-PCR, seroconversion of IgM | VEEV, OROV, GUAV, DENV | 2094 | 16 | 0.8% | unknown | native |

*****HI: hemagglutination inhibition, NT: neutralization test, IA: immunofluorescence assay.

**AURV: Aura virus, CARV: Caraparu virus, CHIKV: chikungunya virus, DENV: dengue virus, EEEV: Eastern equine encephalitis virus, GUAV: Guaroa virus, ILHV: ilheus virus, MADV: Madariaga virus, MUCV: Mucambo virus, OROV: Oropuche virus, ROCV: Rocio virus, SLEV: Saint Louis encephalitis virus, UNAV: Una virus, VEEV: Venezuelan equine encephalitis virus, WEEV: Western equine encephalitis virus, YFV: yellow fever virus, ZIKV: Zika virus.

**S7 Table. Characteristics of MAYV cross-sectional seroprevalence studies in humans.**

| Ref | Study year | Country | State | Diagnostic method* | Total | No. positive | Sero-prevalence | Age stratified | Zone | Indigenous community | Population-based |
| --- | --- | --- | --- | --- | --- | --- | --- | --- | --- | --- | --- |
| [44] | 1997 | Ecuador | Morona-Santiago | ELISA | 91 | 42 | 46.2% | yes | rural | yes, in military service | no |
| [45] | 2007 | Brazil | Amazonia | ELISA | 270 | 119 | 44.1% | yes | rural | no | yes |
| [46] | 1965 | Brazil | Para | HI | 221 | not reported | 37.0% | yes | rural | yes, Gorotire | yes |
| [46] | 1966 | Brazil | Para | HI | 178 | not reported | 47.0% | yes | rural | yes, Tiriyo | yes |
| [46] | 1969 | Brazil | Para | HI | 189 | not reported | 49.0% | yes | rural | yes, Mekranoti | yes |
| [46] | 1970 | Brazil | Para | HI | 69 | not reported | 20.0% | yes | rural | yes, Kuben KK | yes |
| [46] | 1970 | Brazil | Para | HI | 102 | 47 | 46.1% | yes | rural | yes, Xikrin | yes |
| [19] | 1996 | French Guiana |  | HI | 1962 | 124 | 6.3% | yes | rural | no | yes |
| [47] | 1957 | Guyana | Rupununi Savannah | NT | 221 | 126 | 57.0% | yes | rural | yes, Amerindias of the Rupunini | no |
| [47] | 1957 | Trinidad and Tobago |  | NT | 615 | 69 | 11.2% | yes | rural | no | yes |
| [48] | 1960 | Colombia | Santander | NT | 176 | 38 | 21.6% | yes | rural | no | no |
| [49] | 2011 | Peru | Iquitos | IgG ELISA | 70 | 38 | 54.3% | yes | rural | yes, Nueva Esperanza | no |
| [50] | 1966 | Colombia | Amazonas | HI and NT | 396 | 76 | 19.2% | yes | rural | some | no |
| [51] | 1956 | Guyana | Rupununi Savannahs | HI and NT | 176 | 38 | 21.6% | yes | rural | yes, Macusi and wapisiani tribes | no |
| [52] | 1964 | Surinam | Brokopondo | NT | 132 | 88 | 66.7% | yes | rural | no | yes |
| [53] | 1960 | Surinam |  | HI and NT | 500 | 8 | 1.6% | no | rural | no | no |
| [54] | 1965 | Peru |  | HI and NT | 100 | 68 | 68.0% | no | rural | no | no |

*****HI: hemagglutination inhibition, NT: neutralization test, ELISA: enzyme-linked immunosorbent assay.

**S8 Table. Studies with possible evidence of MAYV transmission in humans.**

| Ref | Study years | Country | State | Town/city | Source population | Showed symptoms? | Diagnostic method* | Other arboviruses studied** | Total | No. positive | % positive | Zone | Origin |
| --- | --- | --- | --- | --- | --- | --- | --- | --- | --- | --- | --- | --- | --- |
| [55] | 1999 | Bolivia |  |  | hospital | yes | serology | no | 1 | 1 | 100% | rural | foreign |
| [56] | 1955 | Brazil | Para | Abaetetuba | community | some | NT to SLF | no | 36 | 5 | 13.9% | rural | native |
| [56] | 1955 | Brazil | Para | Altamira | community | some | NT to SLF | no | 24 | 1 | 4.2% | rural | native |
| [56] | 1955 | Brazil | Para | Belterra | community | some | NT to SLF | no | 17 | 5 | 29.4% | rural | native |
| [56] | 1955 | Brazil | Para | Cametá | community | some | NT to SLF | no | 29 | 7 | 24.1% | rural | native |
| [56] | 1955 | Brazil | Para | Capim River | community | some | NT to SLF | no | 33 | 1 | 3.0% | rural | native |
| [56] | 1955 | Brazil | Para | Fordlandia | community | some | NT to SLF | no | 11 | 1 | 9.1% | rural | native |
| [56] | 1955 | Brazil | Amazonas | Labrea | community | some | NT to SLF | no | 24 | 7 | 29.2% | rural | native |
| [56] | 1955 | Brazil | Amazonas | Manaus | community | some | NT to SLF | no | 15 | 1 | 6.7% | rural | native |
| [56] | 1955 | Brazil | Para | Obidos | community | some | NT to SLF | no | 14 | 2 | 14.3% | rural | native |
| [56] | 1955 | Brazil | Para | Belem | community | some | NT to SLF | no | 136 | 3 | 2.2% | rural | native |
| [46] | 1965 | Brazil | Para | Gorotire | community | no | HI | EEEV, MUCV, PIXV, UNAV, AURV, WEEV, YFV, ILHV, BUSV, SLEV, BUNV, MAGV, GUAV | 221 | 90 | 40.7% | rural | native |
| [46] | 1966-1970 | Brazil | Para | Tiriyo | community | no | HI | EEEV, MUCV, PIXV, UNAV, AURV, WEEV, YFV, ILHV, BUSV, SLEV, BUNV, MAGV, GUAV | 217 | 127 | 58.5% | rural | native |
| [57] | 1967 | Brazil | Mato Grosso | Simao Lopes | community | no | HI | ILHV, YFV, BUNV | 155 | 31 | 20.0% | rural | native |
| [57] | 1967 | Brazil | Mato Grosso | Sao Marcos | community | no | HI | ILHV, YFV, BUNV | 257 | 45 | 17.5% | rural | native |
| [46] | 1969-1972 | Brazil | Para | Mekranoti | community | no | HI | EEEV, MUCV, PIXV, UNAV, AURV, WEEV, YFV, ILHV, BUSV, SLEV, BUNV, MAGV, GUAV | 190 | 93 | 48.9% | rural | native |
| [46] | 1970 | Brazil | Para | Kuben Kran Kegn | community | no | HI | EEEV, MUCV, PIXV, UNAV, AURV, WEEV, YFV, ILHV, BUSV, SLEV, BUNV, MAGV, GUAV | 69 | 16 | 23.2% | rural | native |
| [46] | 1970-1972 | Brazil | Para | Krikin | community | no | HI | EEEV, MUCV, PIXV, UNAV, AURV, WEEV, YFV, ILHV, BUSV, SLEV, BUNV, MAGV, GUAV | 102 | 47 | 46.1% | rural | native |
| [58] | 1972 | Brazil | Para | Altamaria | community | yes | HI seroconversion | SELV, WEEV, OROV, GUAV | 832 | 12 | 1.4% | rural | native |
| [59] | 1984 | Brazil | Bahia | Corte de Pedra | hospital | no | HI | WEEV, EEV, MUCV, YFV, BUSV, ILHV, SLEV, Cacipacore virus, ROCV, DENV, Itaporanga virus, Tacaiuma virus, Iaco virus, GUAV, OROV, Utinga virus | 288 | 1 | 0.3% | rural | foreign |
| [60] | 1999-2000 | Brazil | Acre | Rio Branco | community | no | HI seroconversion | MUCV, ILHV, ROCV, DENV, SLEV, YFV, OROV, CARV, Catu virus | 178 | 12 | 6.7% | mixed | native |
| [61] | 2007-2008 | Brazil | Para | Juruti | community | yes | HI | no | 1597 | 20 | 1.3% | rural | native |
| [45] | 2007 | Brazil | Amazonia |  | community | no | IgG ELISA | no | 270 | 119 | 44.1% | rural | native |
| [61] | 2007-2008 | Brazil | Para | Juruti | community | yes | IgM MAC-ELISA | DENV, YFV, OROV | 102 | 5 | 4.9% | rural | native |
| [44] | 1997 | Ecuador | Morona-Santiago |  | community | no | IgG ELISA | no | 91 | 42 | 46.2% | rural | military |
| [19] | 1996 | French Guiania |  |  | community | no | HI | CHIKV | 1305 | 115 | 8.8% | mixed | native |
| [62] | 2010 | Panama | Darien |  | community | some | IgM-ELISA | MADV, VEEV | 72 | 1 | 1.4% | rural | native |
| [63] | 1965 | Peru | Ucayalli | Pucallpa | community | no | HI | AURV, EEEV, MUCV, PIXU, UNAV, VEEV, WEEV | 546 | 164 | 30.0% | mixed | native |
| [63] | 1965 | Peru | Huanuco | TingoMaria | community | no | HI | AURV, EEEV, MUCV, PIXU, UNAV, VEEV, WEEV | 517 | 202 | 39.1% | mixed | native |
| [64] | 2006-2008 | Peru | Loreto | Iquitos | community | no | IgG ELISA | no | 3000 | 486 | 16.2% | rural | native |
| [65] | 2015 | Peru | Loreto | Datem del Marañon | hospital | no | IgG ELISA | DENV, ILHV, SLEV, WNV, YFV, VEEV, UNAV, EEEV, Allpahuayo virus, Tacaribe virus, bunyavirus, CARV, MAGV, MURV, OROV | 364 | 6 | 1.6% | mixed | native |
| [66] | 2017 | Peru | Amazonas |  | community | yes | Culture, IA, RT-PCR | DENV, ZIKV, VEEV, CHIKV, OROV | 1983 | 11 | 0.6% | rural | native |
| [67] | 1961-1962 | Surinam | Marowijne | Albina | community | some | HI | EEEV, DENV, SLEV, ILHV, Cache Valley virus, WEEV, YFV | 340 | 18 | 5.3% | rural | foreign |
| [53] | 1962-1964 | Surinam |  |  | community | some | HI seroconversion | VEEV, UNAV, SLEV, CARV, MUCV, Oriboca virus, Restan virus, Cache Valley virus, Paramaribo virus | 500 | 8 | 1.6% | rural | foreign/  military |

These studies were not classified in other categories but strongly indicate presence of MAYV.

*HI: hemagglutination inhibition, NT: neutralization test, SLF: Semliki Forest virus, IA: immunofluorescence assay.

**AURV: Aura virus, BUNV: Bunyawera virus, BUSV: Bussuquara virus, CARV: Caraparu virus, CHIKV: chikungunya virus, DENV: dengue virus, EEEV: Eastern equine encephalitis virus, GUAV: Guaroa virus, ILHV: ilheus virus, MADV: Madariaga virus, MAGV: Maguari virus, MUCV: Mucambo virus, OROV: Oropuche virus, ROCV: Rocio virus, SLEV: Saint Louis encephalitis virus, UNAV: Una virus, VEEV: Venezuelan equine encephalitis virus, WEEV: Western equine encephalitis virus, YFV: yellow fever virus, ZIKV: Zika virus.

**S9 Table. Studies that detected MAYV in animals.**

| Family | Genus | Species | Year min | Year max | Country | State | Town/ city | Native or migratory birds | Diagnostic method* | Total | No. positive | Antibody titres | Zone | Ref |
| --- | --- | --- | --- | --- | --- | --- | --- | --- | --- | --- | --- | --- | --- | --- |
| Order primates | | | | | | | | | | | | | | |
| Aotidae | *Aotus* |  | 1957 | 1957 | Colombia | Santander | San Vicente de Chucuri |  | HI | 2 | 1 |  | rural | [48] |
| Atelidae | *Alouatta* |  | 1957 | 1957 | Colombia | Llanos Orientales |  |  | HI | 6 | 4 |  | rural | [48] |
| Atelidae | *Alouatta* |  | 1957 | 1957 | Colombia | Santander | San Vicente de Chucuri |  | HI | 5 | 3 |  | rural | [48] |
| Cebidae | *Sapajus* |  | 1957 | 1957 | Colombia | Santander | San Vicente de Chucuri |  | HI | 5 | 1 |  | rural | [48] |
| Icteridae | *Icterus* | *Icterus spurius* | 1967 | 1967 | US | Louisiana |  | migratory | HI, PRNT, viral culture by inoculation into suckling mice | 223 | 1 | yes | rural | [68] |
| Atelidae | *Alouatta* | *Alouatta villosa* | 1974 | 1976 | Panama | Panamá and Darién Provinces | Serranía de Majé |  | PRNT | 5 | 3 |  | sylvatic | [69] |
| Cebidae | *Sapajus* |  | 1978 | 1978 | Brazil | Para | Belterra |  | HI, PNRT | 1 | 1 |  | rural | [70] |
| Callitrichidae |  |  | 1978 | 1978 | Brazil | Para | Belterra |  | HI, PRNT | 119 | 32 |  | rural | [70] |
| Atelidae | *Alouatta* | *Alouatta seniculus* | 1994 | 1995 | French Guiana | Cayenne | Sinnamary |  | HI | 106 | 70 |  | sylvatic | [19] |
| Callitrichidae | *Saguinus* | *Saguinus midas* | 1994 | 1995 | French Guiana | Cayenne | Sinnamary |  | HI | 44 | 8 |  | sylvatic | [19] |
| Atelidae | *Alouatta* | *Alouatta seniculus* | 1994 | 1995 | French Guiana | Cayenne | Sinnamary |  | HI | 98 | 63 |  | sylvatic | [71] |
| Callitrichidae | *Saguinus* | *Saguinus midas* | 1994 | 1995 | French Guiana | Cayenne | Sinnamary |  | HI | 43 | 8 |  | sylvatic | [71] |
| Atelidae | *Alouatta* | *Alouatta seniculus* | 1994 | 1995 | French Guiana | Cayenne | Sinnamary |  | HI, PRNT | 98 | 51 |  | sylvatic | [72] |
| Pitheciidae | *Pithecia* | *Pithecia pithecia* | 1994 | 1995 | French Guiana | Cayenne | Sinnamary |  | HI, PRNT | 5 | 5 |  | sylvatic | [72] |
| Pitheciidae | *Pithecia* | *Pithecia pithecia* | 1994 | 1995 | French Guiana | Cayenne | Sinnamary |  | HI, PRNT | 5 | 4 |  | sylvatic | [72] |
| Cebidae | *Saimiri* | *Saimiri sciureus* | 1994 | 1995 | French Guiana | Cayenne | Sinnamary |  | HI, PRNT | 6 | 4 |  | sylvatic | [72] |
| Callitrichidae | *Saguinus* | *Saguinus midas* | 1994 | 1995 | French Guiana | Cayenne | Sinnamary |  | HI, PRNT | 42 | 8 |  | sylvatic | [72] |
| Atelidae | *Lagothrix* | *Lagothrix poeppigii* | 2007 | 2008 | Peru | Loreto | Maynas |  | ELISA, PRNT | 11 | 6 |  | rural | [49] |
| Atelidae | *Alouatta* | *Alouatta seniculus* | 2007 | 2008 | Peru | Loreto | Maynas |  | ELISA, PRNT | 1 | 1 |  | rural | [49] |
| Pitheciidae | *Cacajao* | *Cacajao calvus* | 2007 | 2008 | Peru | Loreto | Maynas |  | ELISA, PRNT | 3 | 1 |  | rural | [49] |
| Cebidae | *Sapajus* | *Sapajus macrocephalus* | 2007 | 2008 | Peru | Loreto | Maynas |  | ELISA, PRNT | 6 | 1 |  | rural | [49] |
| Cebidae | *Sapajus* | *Sapajus libidinosus* | 2008 | 2010 | Brazil | Alagoas | Maceió |  | HI | 5 | 1 | yes | captivity | [73] |
| Cebidae | *Sapajus* | *Sapajus libidinosus* | 2008 | 2010 | Brazil | Paraíba | Cabedelo |  | HI | 37 | 12 | yes | captivity | [73] |
| Cebidae | *Sapajus* | *Sapajus libidinosus* | 2008 | 2010 | Brazil | Pernambuco | Recife |  | HI | 16 | 4 | yes | captivity | [73] |
| Cebidae | *Sapajus* | *Sapajus libidinosus* | 2008 | 2010 | Brazil | Rio Grande do Norte | Natal |  | HI | 16 | 7 | yes | captivity | [73] |
| Cebidae | *Sapajus* | *Sapajus libidinosus* | 2008 | 2010 | Brazil | Piauí | Teresina |  | HI | 26 | 5 | yes | captivity | [73] |
| Cebidae | *Sapajus* | *Cebus apella* | 2009 | 2010 | Brazil | Mato Grosso do Sul | Bonito |  | HI | 35 | 7 | yes | sylvatic | [74] |
| Atelidae | *Alouatta* | *Alouatta caraya* | 2010 | 2010 | Brazil | Mato Grosso do Sul | Campo Grande |  | HI | 2 | 2 | yes | sylvatic | [74] |
| Atelidae | *Ateles* | *Ateles marginatus* | 2012 | 2017 | Brazil | Bahia | Salvador |  | HI, PRNT | 3 | 1 | yes | captivity | [75] |
| Cebidae | *Sapajus* | *Sapajus xanthosternos* | 2012 | 2017 | Brazil | Bahia | Salvador |  | HI, PRNT | 11 | 1 | yes | captivity | [75] |
| Cebidae | *Sapajus spp* |  | 2013 | 2013 | Brazil | Mato Grosso do Sul | Jardim |  | HI | 13 | 1 | yes | sylvatic | [76] |
| Order Rodentia | | | | | | | | | | | | | | |
| Dasyproctidae | *Dasyprocta* | *Dasyprocta punctata* | 1974 | 1976 | Panama | Provincia de Panamá y Provincia de Darién | Serranía de Majé |  | PRNT | 5 | 3 |  | sylvatic | [69] |
| Dasyproctidae | *Dasyprocta* | *Dasyprocta leporina* | 1994 | 1995 | French Guiana | Cayenne | Sinnamary |  | HI, PRNT | 29 | 5 |  | sylvatic | [72] |
| Erethizontidae | *Coendou* | *Coendou prehensilis* | 1994 | 1995 | French Guiana | Cayenne | Sinnamary |  | HI, PRNT | 26 | 3 |  | sylvatic | [72] |
| Erethizontidae | *Coendou* | *Coendou melanurus* | 1994 | 1995 | French Guiana | Cayenne | Sinnamary |  | HI, PRNT | 15 | 2 |  | sylvatic | [72] |
| Echimyidae | *Proechimys* |  | 1994 | 1995 | French Guiana | Cayenne | Sinnamary |  | HI, PRNT | 18 | 1 |  | sylvatic | [72] |
| Echimyidae | *Echimys* |  | 1994 | 1995 | French Guiana | Cayenne | Sinnamary |  | HI, PRNT | 21 | 1 |  | sylvatic | [72] |
| Dasyproctidae | *Dasyprocta* | *Dasyprocta fuliginosa* | 2007 | 2008 | Peru | Loreto | Maynas |  | ELISA, PRNT | 27 | 3 |  | rural | [49] |
| Cuniculidae | *Cuniculus* | *Cuniculus paca* | 2007 | 2008 | Peru | Loreto | Maynas |  | ELISA, PRNT | 10 | 1 |  | rural | [49] |
| Order Columbiformes | | | | | | | | | | | | | | |
| Columbidae |  |  | 1978 | 1978 | Brazil | Para | Belterra | native | HI, PRNT | 34 | 1 |  | rural | [70] |
| Order Caprimulgiformes | | | | | | | | | | | | | | |
| Caprimulgidae |  |  | 1978 | 1978 | Brazil | Para | Belterra | native | HI, PRNT | 5 | 1 |  | rural | [70] |
| Order Passeriformes | | | | | | | | | | | | | | |
| Dendrocolaptiae |  |  | 1978 | 1978 | Brazil | Para | Belterrra | native | HI, PRNT | 97 | 1 |  | rural | [70] |
| Formicariidae |  |  | 1978 | 1978 | Brazil | Para | Belterrra | native | HI, PRNT | 444 | 5 |  | rural | [70] |
| Pipridae |  |  | 1978 | 1978 | Brazil | Para | Belterrra | native | HI, PRNT | 229 | 1 |  | rural | [70] |
| Tyrannidae |  |  | 1978 | 1978 | Brazil | Para | Belterrra | migratory | HI, PRNT | 102 | 1 |  | rural | [70] |
| Fringillidae |  |  | 1978 | 1978 | Brazil | Para | Belterrra |  | HI, PRNT | 131 | 6 |  | rural | [70] |
| Order Pilosa | | | | | | | | | | | | | | |
| Megalonychidae | *Choloepus* | *Choloepus didactylus* | 1994 | 1995 | French Guiana | Cayenne | Sinnamary |  | HI, PRNT | 26 | 7 |  | sylvatic | [72] |
| Bradypodidae | *Bradypus* | *Bradypus tridactylus* | 1994 | 1995 | French Guiana | Cayenne | Sinnamary |  | HI, PRNT | 29 | 1 |  | sylvatic | [72] |
| Myrmecophagidae | *Tamandua* | *Tamandua tetradactyla* | 1994 | 1995 | French Guiana | Cayenne | Sinnamary |  | HI, PRNT | 26 | 6 |  | sylvatic | [72] |
| Order Cingulata | | | | | | | | | | | | | | |
| Dasypodidae | *Dasypus* | *Dasypus novemcinctus* | 1994 | 1995 | French Guiana | Cayenne | Sinnamary |  | HI, PRNT | 40 | 4 |  | sylvatic | [72] |
| Didelphidae | *Didelphis* | *Didelphis marsupialis* | 1994 | 1995 | French Guiana | Cayenne | Sinnamary |  | HI, PRNT | 29 | 1 |  | sylvatic | [72] |
| Dasypodidae | *Dasypus* | *Dasypus novemcinctus* | 2007 | 2008 | Peru | Loreto | Maynas |  | ELISA, PRNT | 4 | 2 |  | rural | [49] |
| Order Didelphimorphia | | | | | | | | | | | | | | |
| Didelphidae | *Didelphis* | *Didelphis albiventris* | 1994 | 1995 | French Guiana | Cayenne | Sinnamary |  | HI, PRNT | 19 | 2 |  | sylvatic | [72] |
| Didelphidae | *Philander* | *Philander opossum* | 1994 | 1995 | French Guiana | Cayenne | Sinnamary |  | HI, PRNT | 27 | 5 |  | sylvatic | [72] |
| Didelphidae | *Caluromys* | *Caluromys philander* | 1994 | 1995 | French Guiana | Cayenne | Sinnamary |  | HI, PRNT | 5 | 1 |  | sylvatic | [72] |
| Order Carnivora | | | | | | | | | | | | | | |
| Procyonidae | *Potos* | *Potos flavus* | 1994 | 1995 | French Guiana | Cayenne | Sinnamary |  | HI, PRNT | 9 | 1 |  | sylvatic | [72] |
| Mustelidae | *Eira* | *Eira barbara* | 1994 | 1995 | French Guiana | Cayenne | Sinnamary |  | HI, PRNT | 7 | 1 |  | sylvatic | [72] |
| Order Artiodactyla | | | | | | | | | | | | | | |
| Tayassuidae | *Pecari* | *Pecari tajacu* | 2007 | 2008 | Peru | Loreto | Maynas |  | ELISA, PRNT | 6 | 1 |  | rural | [49] |
| Order Perissodactyla | | | | | | | | | | | | | | |
| Equidae |  |  | 2009 | 2010 | Brazil | Mato Grosso do Sul | Pantanal |  | HI, PRNT | 748 | 44 | yes | rural | [77] |
| Order Crocodilia | | | | | | | | | | | | | | |
| Alligatoridae |  |  | 2009 | 2010 | Brazil | Mato Grosso do Sul | Pantanal |  | HI, PRNT | 87 | 2 | yes | sylvatic | [77] |

*****HI: hemagglutination inhibition, PRNT: plaque reduction neutralization test, ELISA: enzyme-linked immunosorbent assay.

**Supplementary results from phylogenetic analysis**

**Sequence selection and alignment**

We screened GenBank on November 15, 2019 for all published sequences of MAYV. We selected only complete or near complete genomes (> 11000 nt). Sixty-nine sequences were identified but four duplicates were excluded (TRVL 15537 KP842810 – FPY0046 KP842813 – BeAr20290 KT754168 – FPI0179 KP842816). The geographic distribution of these sequences was from Bolivia (n= 6), Brazil (n = 29), French Guiana (n=2), Haiti (n= 5), Peru (n = 17) Trinidad and Tobago (n = 2), and Venezuela (n = 7). Location, source, and date of sample were retrieved from GenBank or original publications. Complete genomes were sampled between 1954 and 2015 (S10 Table). Sequences were aligned using MUSCLE algorithm [78] with MEGA7 software [79] and manually edited to maintain codon homology.

**Phylogenetic signal and maximum-likelihood phylogeny inference**

We evaluated the substitution saturation by Xia test in DAMBE7 [80]. Full-genome analysis has a low saturation, meaning the sequences are sufficient to identify phylogenetic signal (Num OTU 32, Iss 0.064 vs Iss.cAsym 0.572, p < 0.001). The best-fitting nucleotide substitution model was selected with jModelTest 2 software [81, 82] according to Bayesian information criterion (BIC). The maximum likelihood tree was constructed by generalized time-reversible + invariable sites + gamma 4 model (S11 Table) with IQ-TREE software**.** The statistical robustness of the tree topology was calculated with Ultrafast bootstrap support and Shimodaira-Hasegawa-like approximate ratio test (SH-aLRT) with 2000 replicates were used to assess the statistical robustness of topologies for internal branching [83, 84]. Strong statistical support along the branches was defined: BS > 75 and/or SH-aLRT > 95.

**S10 Table.** **Full genomes of MAYV included in the phylogenetic analysis.**

| GenBank number accession | Internal ID | Strain | Length bp | Country | State | Town / city | Year of collection | Source |
| --- | --- | --- | --- | --- | --- | --- | --- | --- |
| MK573246 | BOL_Uruma_1955 | Uruma | 11206 | Bolivia | Santa Cruz | Santa Cruz | 1955 | human |
| MK573245 | BOL_FSB0311_2002 | FSB0311 | 11206 | Bolivia |  |  | 2002 | human |
| KP842817 | BOL_FVB0069_2006 | FVB0069 | 11093 | Bolivia |  |  | 2006 | human |
| KP842814 | BOL_FVB0112_2006 | FVB0112 | 11099 | Bolivia |  |  | 2006 | human |
| KP842806 | BOL_FSB1131_2006 | FSB1131 | 11099 | Bolivia |  |  | 2006 | human |
| KP842805 | BOL_FSB0319_2002 | FSB0319 | 11099 | Bolivia |  |  | 2002 | human |
| MK573244 | BRA_BeH343155_1978 | BeH343155 | 11206 | Brazil |  |  | 1978 | human |
| MK573241 | BRA_BeH506151_1991 | BeH506151 | 11206 | Brazil |  |  | 1991 | human |
| MK573239 | BRA_BeH428890_1984 | BeH428890 | 11224 | Brazil |  |  | 1984 | human |
| MK573238 | BRA_BeH407_1955 | BeH407 | 11224 | Brazil |  |  | 1955 | human |
| KY618140 | BRA_BeH792430_2012 | BeH792430 | 11480 | Brazil | Para | Barcarena | 2012 | human |
| KY618139 | BRA_BeH758762_2012 | BeH758762 | 11365 | Brazil | Para | Parauapebas | 2009 | human |
| KY618138 | BRA_BeH744173_2008 | BeH744173 | 11381 | Brazil | Para | Santa Barbara do Para | 2008 | human |
| KY618137 | BRA_BeH744141_2008 | BeH744141 | 11381 | Brazil | Para | Belem | 2008 | human |
| KY618136 | BRA_BeH743921_1991 | BeH743921 | 11381 | Brazil | Para | Santa Barbara do Para | 2008 | human |
| KY618135 | BRA_BeH505465_1991 | BeH505465 | 11612 | Brazil | Para | Belem | 1991 | human |
| KY618134 | BRA_BeH504639_1991 | BeH504639 | 11512 | Brazil | Goias | Goiania | 1991 | human |
| KY618133 | BRA_BeH473130_1988 | BeH473130 | 11535 | Brazil | Para | Santarem | 1988 | human |
| KY618132 | BRA_BeH394885_1981 | BeH394885 | 11416 | Brazil | Para | Barro Branco | 1981 | human |
| KY618131 | BRA_BeH342916_1978 | BeH342916 | 11423 | Brazil | Para | Santarem | 1978 | human |
| KY618130 | BRA_BeAr757954_2011 | BeAr757954 | 11550 | Brazil | Rio Grande do Sul |  | 2011 | *Culex* sp. |
| KY618129 | BRA_BeAr505578_1991 | BeAr505578 | 11541 | Brazil | Para | Benevides | 1991 | *Haemagogus janthinomys* |
| KY618128 | BRA_BeAr344910_1978 | BeAr344910 | 11407 | Brazil | Para | Santarem | 1978 | *Haemagogus janthinomys* |
| KY618127 | BRA_BeAr20290_1960 | BeAr20290 | 11472 | Brazil |  |  | 1960 | *Haemagogus* sp. |
| KT818520 | BRA_LPV01_2014 | BR/SJRP/LPV01/2015 | 11438 | Brazil |  |  | 2014 | human |
| KP842820 | BRA_BeAr30853_1961 | BeAr30853 | 11105 | Brazil | Para |  | 1961 | *Ixodes* sp*.* |
| KP842819 | BRA_BeH256_1955 | BeH256 | 11105 | Brazil |  |  | 1955 | human |
| KP842818 | BRA_BeAr505411_1991 | BeAr505411 | 11141 | Brazil |  |  | 1991 | *Haemagogus janthinomys* |
| KP842809 | BRA_BeH186258_1970 | BeH186258 | 11099 | Brazil |  |  | 1970 | human |
| KP842804 | BRA_BeAn337622_1978 | BeAn337622 | 11099 | Brazil |  |  | 1978 | human |
| KP842803 | BRA_BeH343148_1978 | BeH343148 | 11099 | Brazil |  |  | 1978 | human |
| KP842802 | BRA_BeAn343102_1978 | BeAn343102 | 11099 | Brazil |  |  | 1978 | monkey |
| KM400591 | BRA_Acre27_2004 | Acre27 | 11273 | Brazil | Acre | Acrelandia | 2004 | human |
| MH513597 | BRA_H307_2015 | BR/Sinop/H307/2015 | 11147 | Brazil | Matto Grosso | Sinop | 2015 | human |
| KJ013266 | FRG_BNI-1_2013 | BNI-1 | 11376 | French Guiana |  |  | 2013 | human |
| DQ001069 | FRG_MAYLC | MAYLC | 11429 | French Guiana |  |  | 1996 | human |
| MK837007 | HAI_0737_2014 | Homo sapiens/Haiti-0737/2014 | 11429 | Haiti | Oest | Port-au-Prince | 2014 | human |
| MK837006 | HAI_0380_2014 | Homo sapiens/Haiti-0380/2014 | 11429 | Haiti | Oest | Port-au-Prince | 2014 | human |
| MN138459 | HAI_0729_2014 | Homo sapiens/Haiti-0729/2014 | 11429 | Haiti | Oest | Port-au-Prince | 2014 | human |
| KY985361 | HAI_1_2014 | Homo sapiens/Haiti-1/2014 | 11462 | Haiti |  |  | 2014 | human |
| KX496990 | HAI_1_2015 | Homo sapiens/Haiti-1/2015 | 11462 | Haiti |  |  | 2015 | human |
| MK573243 | PER_IQU2950_2000 | IQU2950 | 11206 | Peru | Loreto | Iquitos | 2000 | human |
| MK573242 | PER_OBS2209_1955 | OBS2209 | 11206 | Peru |  |  | 1995 | human |
| MK070491 | PER_IQT4235_1997 | IQT4235 | 11413 | Peru |  |  | 1997 | human |
| KY026200 | PER_FPI179_2011 | FPI0179 | 11093 | Peru |  |  | 2011 | human |
| KY026199 | PER_FPY0122_2011 | FPY_0122 | 11443 | Peru |  |  | 2011 | human |
| KY026198 | PER_FPY0046_2011 | FPY0046 | 11099 | Peru |  |  | 2011 | human |
| KY026197 | PER_FPI1738_2011 | FPI_1738 | 11438 | Peru |  |  | 2011 | human |
| KY026195 | PER_FPI1766_2011 | FPI_1766 | 11456 | Peru |  |  | 2011 | human |
| KP842816 | PER_FPI0179_2011 | FPI0179 | 11093 | Peru |  |  | 2011 | human |
| KP842815 | PER_FPI1761_2011 | FPI1761 | 11093 | Peru |  |  | 2011 | human |
| KP842813 | PER_FPY0046_2011 | FPY0046 | 11099 | Peru |  |  | 2011 | human |
| KP842812 | PER_FMD3213_2010 | FMD3213 | 11099 | Peru |  |  | 2010 | human |
| KP842811 | PER_FMD0641_2005 | FMD0641 | 11099 | Peru |  |  | 2005 | human |
| KP842808 | PER_IQU3056_2000 | IQU3056 | 11099 | Peru |  |  | 2000 | human |
| KP842807 | PER_Ohio_1995 | Ohio | 11099 | Peru |  |  | 1995 | human |
| KP842801 | PER_IQE2777_2006 | IQE2777 | 11099 | Peru |  |  | 2006 | human |
| KP842800 | PER_ARV0565_1995 | ARV0565 | 11099 | Peru |  |  | 1995 | human |
| MK573240 | TRI_TRVL15537_1957 | TRVL15537 | 11202 | Trinidad and Tobago |  |  | 1957 | *Manzonia venezuelensis* |
| MK070492 | TRI_TRVL 4675_1954 | TRVL 4675 | 11424 | Trinidad and Tobago |  |  | 1954 | human |
| MK288026 | VEN_1_2016 | Homo sapiens/Venezuela-1/2016 | 11441 | Venezuela | Portuguesa | Ospino | 2016 | human |
| KP842799 | VEN_MAYV15A_2010 | MAYV15A | 11099 | Venezuela | Portuguesa | Ospino | 2010 | human |
| KP842798 | VEN_MAYV14A_2010 | MAYV14A | 11099 | Venezuela | Portuguesa | Ospino | 2010 | human |
| KP842797 | VEN_MAYV13A_2010 | MAYV13A | 11099 | Venezuela | Portuguesa | Ospino | 2010 | human |
| KP842796 | VEN_MAYV12A_2010 | MAYV12A | 11099 | Venezuela | Portuguesa | Ospino | 2010 | human |
| KP842795 | VEN_MAYV11A_2010 | MAYV11A | 11099 | Venezuela | Portuguesa | Ospino | 2010 | human |
| KP842794 | VEN_MAYV16A_2010 | MAYV16A | 11099 | Venezuela | Portuguesa | Ospino | 2010 | human |

**S11 Table. Nucleotide substitution models.** The best-fitting model is in bold.

| Model | Number of estimated parameters | Log likelihood | BIC |
| --- | --- | --- | --- |
| GTR+I+G | **138** | **-38644.6** | **78574** |
| GTR+G | 137 | -38652.8 | 78581 |
| GTR+I | 137 | -38668.8 | 78613 |
| SYM+I+G | 135 | -38687.9 | 78632 |
| SYM+G | 134 | -39695.6 | 78639 |
| SYM+I | 134 | -38712.3 | 78672 |
| TrN+I+G | 135 | -38717.8 | 78692 |
| TrN+G | 134 | -38725.5 | 78698 |
| TIM1+I+G | 136 | -38717.7 | 78701 |
